# Impact of B.1.351 (beta) SARS-CoV-2 variant on BNT162b2 mRNA vaccine effectiveness in long-term care facilities of eastern France: a retrospective cohort study

**DOI:** 10.1101/2021.07.28.21261285

**Authors:** Benjamin Lefèvre, Laura Tondeur, Yoann Madec, Rebecca Grant, Bruno Lina, Sylvie van der Werf, Christian Rabaud, Arnaud Fontanet, Antoine Legoff

## Abstract

**Background:** We aimed to assess the effectiveness of the BNT162b2 mRNA vaccine against B.1.351 (beta) variant among residents of long-term care facilities (LCTFs) in eastern France.

**Methods:** We used routinely collected surveillance and COVID-19 vaccination data to conduct a retrospective cohort study of SARS-CoV-2 B.1.351 infection incidence and vaccine effectiveness among LCTFs residents in eastern France between 15 January and 19 May 2021. Data from secondary RT-PCR screening were used to identify B.1.351 variants.

**Findings:** Included in our analysis were 378 residents from five LCTFs: 287 (76%) females, with median (IQR) age of 89 (83-92) years. Two B.1.351 outbreaks took place in LTCFs in which more than 70% of residents had received two doses of BNT162b2 mRNA vaccine, which included 11 cases of severe disease and six deaths among those who had received two doses. Vaccine effectiveness (95% CI) seven days after the second dose of vaccine was 49% (14-69) against any infection with B.1.351 and 86% (67-94) against severe forms of COVID-19. In multivariable analysis, females were less likely to develop severe forms of disease (IRR = 0.35, 95% CI = 0.20-0.63).

**Interpretation:** We observed reduced vaccine effectiveness associated with B.1.351, as well as B.1.351 outbreaks in two LTCFs among individuals who had received two doses of vaccine. Our findings highlight the need to maintain SARS-CoV-2 surveillance in these high-risk settings beyond the current COVID-19 mass vaccination campaign, and advocate for a booster vaccine dose prior to the next winter season.

## Introduction

In late 2020, England experienced a resurgence in incidence of SARS-CoV-2 which was later attributed to the emergence of the SARS-CoV-2 B.1.1.7 (alpha) variant – a variant which has demonstrated increased transmissibility compared to pre-existing original European lineages such as 20A and 20E (EU)^1^. This was followed by the emergence of the B.1.351 (beta) variant in South Africa and the P.1 (gamma) variant in Brazil, both of which emerged in a context of rapid resurgence in SARS-CoV-2 incidence^2,3^.

Of further concern from a public health perspective are the mutations in the SARS-CoV-2 Spike protein in the B.1.351 (E484K and K417N) and P.1 (E484K and K417T) lineages with potential escape to SARS-CoV-2 antibodies. The potential for immune escape has been investigated through evaluation of the neutralizing capacity of sera or plasma from individuals with past infection against B.1.351 and P.1^4-7^, and sera from individuals having received mRNA COVID-19 vaccines against B.1.351 and P.1^7-9^. Overall, the neutralizing activity seems to be similar for the original non-variant viruses and B.1.1.7, reduced for B.1.351, and intermediate for P.1.

Real-world evaluations of COVID-19 vaccines are available against the original non-variant viruses and B.1.1.7, showing vaccine effectiveness to be 85-94% against symptomatic infection beyond seven days after the second dose of BNT162b2 mRNA COVID-19 vaccine^10,11^. For B.1.351, lower efficacy of the ChAdOx1 nCoV-19^12^, Ad26.COV2.S^13^, and NVX-CoV2372^14^ vaccines have been shown during clinical trials in South Africa and breakthrough infections with B.1.1.7 and B.1.351 variants have been documented in individuals vaccinated with BNT162b2 mRNA vaccine in Israel^15^. A recent publication estimated the effectiveness of two doses of BNT162b2 mRNA vaccine to be 75% against all clinical forms of B.1.351 infection, and 97% against severe, critical or fatal disease in Qatar^16^.

The COVID-19 vaccination program began in France on 27 December 2020, with early prioritization of BNT162b2 mRNA vaccine for residents of long-term care facilities (LTCFs) from early January 2021.^17^ Periodic nationwide whole genome sequencing for the surveillance of SARS-CoV-2 variants began in the third week of January 2021, with high levels of circulation of B.1.351 observed in eastern France in early February.^18^ In Moselle, daily screening of variants from positive RT-PCR test results showed a rise in the proportion of B.1.351 with peak of 47% of all positive RT-PCR test results on 28 February 2021, followed by a gradual decline to 11% on 16 May 2021.^19^ During this time, several COVID-19 outbreaks in LTCFs in the same region were notified, despite the vaccination campaign and the prioritization of residents for vaccination.

Here, we use surveillance data routinely collected in LTCFs in eastern France to estimate the vaccine effectiveness of BNT162b2 mRNA vaccine against B.1.351 among residents of LTCFs.

## Methods

### Study design and participants

In LTCF, SARS-CoV-2 surveillance is organized through RT-PCR testing of nasopharyngeal swabs whenever a resident or a health care worker (HCW) has symptoms suggestive of COVID-19. In addition, HCWs are asked to be tested on a weekly and voluntary basis, with all residents tested in the event that a HCW returns a positive test. Since early January 2021, all LTCF residents have been offered vaccination with BNT162b2 mRNA vaccine. HCWs working in LTCFs have also been prioritized for vaccination. At the beginning of the vaccination campaigns in LTCFs, both for residents and HCWs, the recommended interval between doses of BNT162b2 mRNA vaccine was 3 weeks.^17^

### Data collection

For this study, we reviewed surveillance data from all LTCFs in three departments (geographical administrative unit) in eastern France where B.1.351 has been circulating since early February 2021. We selected LTCFs in which any SARS-CoV-2 outbreak that implicated B.1.351 had been documented between 15 January and 16 April 2021. In each LTCF, all residents were eligible for inclusion and the surveillance data collected by medical personnel included age, sex, history of RT-PCR positive SARS-CoV-2 infection prior to 15 January 2021, history of COVID-19 vaccination, and history of positive RT-PCR for SARS-CoV-2 infection since 15 January 2021. SARS-CoV-2 infections were categorized as mild if the resident had no symptoms or symptoms that did not require oxygen support and remained in the facility; severe if the resident had symptoms that required oxygen support and/or the resident was transferred to a hospital, or the resident died. These data were used to build a retrospective cohort of LTCF residents from 15 January 2021 to 19 May 2021.

### Identification of SARS-CoV-2 B.1.351

During the third week of January 2021, a second round of RT-PCR called screening RT-PCR was implemented in France to identify SARS-CoV-2 variants of concern (VOC). This screening strategy focused on the detection of the B.1.1.7, B.1.351 and P.1 variants.^20^ Screening relied on the detection of the N501Y mutation shared by these three VOCs and one or two additional targets specific of either the B.1.1.7 lineage (del69-70, A570D, P681H) or the B.1.351 and/or P.1 lineages (K417N, E484K). When screening RT-PCR was negative for all targets, the virus was assumed to belong to a pre-existing original European non-variant lineages.

During outbreaks in LTCFs in eastern France, not all samples underwent secondary RT-PCR screening for identification of variants. For the purpose of this analysis, the available secondary screening data were used to infer the virus circulating in the LTCF during the outbreak. Since all RT-PCR screened viruses of the same cluster belonged to the same lineage, the remaining unscreened viruses of the same cluster were considered to belong to the same lineage. As nationwide surveillance of variants indicated that the P.1 lineage was not circulating in eastern France at the time of the study, all targets identifying B.1.351/P.1 lineages (K417N, E484K) were considered to be B.1.351.^21^

### Statistical analyses

The primary objective of the analysis was to determine the vaccine effectiveness fourteen days after the first dose, and seven days after the second dose of BNT162b2 mRNA vaccine against B.1.351 infection and severe disease. Seven days after the second dose of BNT162b2 mRNA vaccine was used for comparability with recently published vaccine effectiveness studies of the same vaccine.^10,11^ Participants contributed person-time from 15 January 2021 onwards, until either the participant tested positive by RT-PCR for SARS-CoV-2 infection, or the end of data collection in the LTCF (which ranged from 29 March to 19 May 2021). Vaccination status was considered as a time-dependent variable, with each participant contributing person-time as non-vaccinated until thirteen days after the first dose, as vaccinated with one dose of vaccine until six days after the second dose, and as vaccinated with two doses of vaccines from seven days after the second dose. Incidence rates (IRs) and incidence rate ratios (IRRs), and their 95% confidence intervals (95% CI), were calculated assuming a Poisson distribution of events. A random effect was added to the model to account for any ‘centre effect’ of each of the facilities. Factors evaluated for association with SARS-CoV-2 infection were age, gender, calendar week, history of past SARS-CoV-2 infection, and COVID-19 vaccination status. The vaccine effectiveness was calculated as one minus the adjusted IRR. All statistical analyses were performed using Stata 16.0 (StataCorp, College Station, TX, USA).

### Ethical considerations

In France, surveillance of COVID-19 testing and COVID-19 vaccination data in LTCFs is considered as public health surveillance which does not require formal ethical review. The pseudonymized data collection by medical personnel at the LTCFs and analysis by the authors have been performed under the legal responsibility of the Société de Pathologie Infectieuse de Langue Française, which ensured compliance with data protection regulations in France.

## Results

Between 29 March and 19 May 2021, five LTCFs were recruited in the study. Facilities ranged in size from 42 to 100 residents, giving a total study population of 378 residents. Of these, 287 (75.9%) were female, and the median (interquartile range (IQR)) age was 89 (83-92) years. The proportion of residents with SARS-CoV-2 infection prior to 15 January 2021 was 20/378 (5.3%), of which six (30.0%) were recent (<3 months), and five (25.0%) were considered clinically severe. By the end of the study period, 279 (73.8%) residents had received two doses of BNT162b2 mRNA vaccine, 59 (15.6%) had received one dose, and 40 (10.6%) had not received any dose. The proportion with past infection was not different between those who had received at least one vaccine dose and those who had not received any vaccine dose (5.3% and 5.0%, respectively).

Overall, there were 145 (38.4%) study participants who were infected during the study period. Of the 145 infections, 53 (36.6%) were severe, including 37 deaths. All infections took place during outbreaks, except for two which were isolated, and for which screening RT-PCR test results were missing (n=1) or undetermined (n=1). Table 1 describes the characteristics of the residents by RT-PCR test result and by severity of infection, excluding the two residents whose infections were isolated.

**Table 1.** Characteristics of long-term care facility residents by RT-PCR test results and clinical forms of infection (n=376)* included in the retrospective cohort study from 15 January to 19 May 2021

|  | PCR-negative<br>(N=233)<br>n (%) | B.1.351 infection |  |
| --- | --- | --- | --- |
|  |  | Mild<br>(N=91)<br>n (%) | Severe<br>(N=52)<br>n (%) |
| Sex |  |  |  |
| Male | 50 (21.5) | 18 (19.8) | 23 (44.2) |
| Female | 183 (78.5) | 73 (80.2) | 29 (55.8) |
| Age (in years) |  |  |  |
| 55-84 | 68 (29.2) | 33 (36.3) | 13 (25.0) |
| 85-94 | 136 (58.4) | 46 (50.5) | 36 (69.2) |
| 94-104 | 29 (12.4) | 12 (13.2) | 3 (5.8) |
| History of past SARS-CoV-2 infection |  |  |  |
| None | 214 (91.9) | 91 (100.0) | 52 (100.0) |
| Asymptomatic | 5 (2.1) | 0 (0.0) | 0 (0.0) |
| Mild | 7 (3.0) | 0 (0.0) | 0 (0.0) |
| Severe | 5 (2.1) | 0 (0.0) | 0 (0.0) |
| Unknown | 2 (0.9) |  |  |
| Long-term care facility |  |  |  |
| A | 30 (12.9) | 18 (19.8) | 23 (44.2) |
| B | 59 (25.3) | 7 (7.7) | 2 (3.9) |
| C | 21 (9.0) | 17 (18.7) | 4 (7.7) |
| D | 61 (26.2) | 25 (27.4) | 10 (19.2) |
| E | 62 (26.6) | 24 (26.4) | 13 (25.0) |
| BNT162b2 mRNA COVID-19 vaccine received |  |  |  |
| None | 22 (9.5) | 5 (5.5) | 13 (25.0) |
| One dose | 18 (7.7) | 17 (18.7) | 24 (46.1) |
| Two doses | 193 (82.8) | 69 (75.8) | 15 (28.9) |

\*Two participants with positive RT-PCR test results, but variant screening RT-PCR test results missing (n=1) or undetermined (n=1), and not part of a cluster, are not shown

Figure 1 shows the timing of the outbreaks and of the vaccination campaigns. In two LCTF (D and E), the outbreak started four and six weeks after the second dose of the vaccination campaign, respectively. LTCF D had 97 residents, among which 70 (72%) had received two doses of vaccine at the time of the outbreak: 35 were infected during the outbreak, including 26 who were infected more than seven days after receiving the second dose of vaccine; among these 26, four had severe disease, including two deaths. LTCF E had 100 residents, among which 78 (78%) had received two doses of vaccine at the time of outbreak: 37 were infected, including 28 who were infected more than seven days after receiving the second dose of vaccine; among these 28, seven had severe disease, including four deaths. In this facility, 17 of the 20 RT-PCR-positive screened as B.1.351/P.1 variant were sequenced and confirmed as B.1.351.

**Figure 1:**
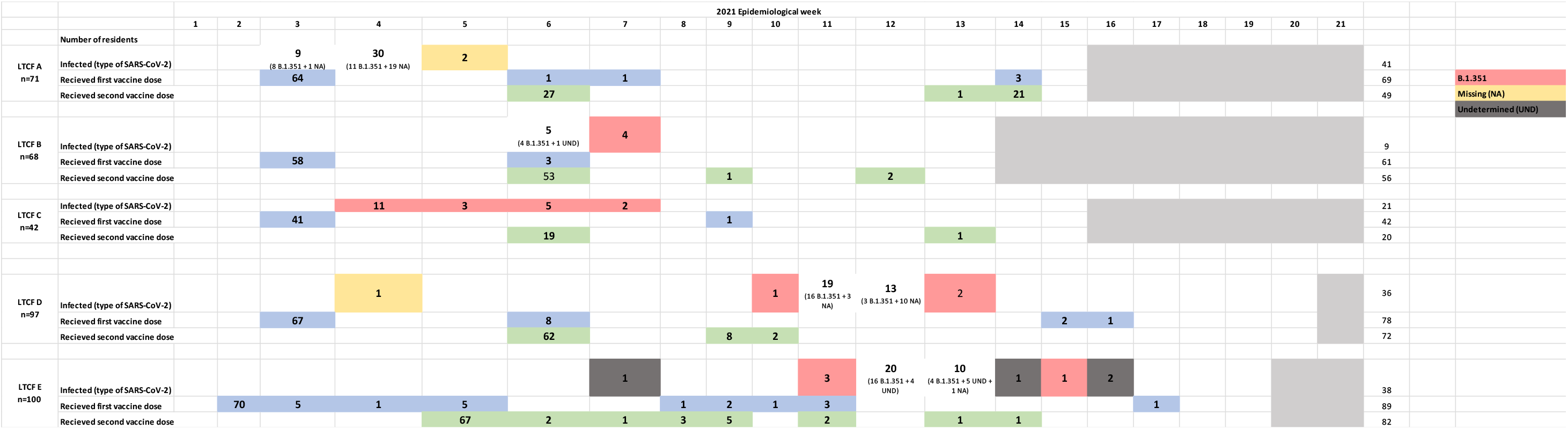
Timing of BNT162b2 mRNA COVID-19 vaccination and B.1.351 outbreaks in LCTFs of eastern France.

Table 2 displays the IRR of the association between characteristics of LTCF residents and B.1.351 infection. A history of past infection was fully protective (IRR = 0; 95% CI = 0.00-0.41), but there were only a limited number of participants in this category. Seven days after the second dose of BNT162b2 mRNA COVID-19 vaccine, the vaccine effectiveness against all forms of infection was estimated at 49% (14-69).

**Table 2.** IRR of the association between characteristics of LTCF residents and B.1.351 infection

|  | Person years<br>(N=81.6) | N=143 | IRR<br>(95% CI) | IRR<sub>a</sub>*<br>(95% CI) |
| --- | --- | --- | --- | --- |
| Sex |  |  |  |  |
| Male | 18.7 | 41 | 1 | 1 |
| Female | 62.9 | 102 | 0.71 (0.49-1.02) | 0.73 (0.50-1.06) |
| Age (in years) |  |  |  |  |
| 55-84 | 24.5 | 46 | 1 | 1 |
| 85-94 | 47.2 | 82 | 0.85 (0.59-1.22) | 0.86 (0.59-1.25) |
| 94-104 | 9.9 | 15 | 0.82 (0.46-1.47) | 0.79 (0.44-1.43) |
| History of past SARS-CoV-2 infection |  |  |  |  |
| No | 76.7 | 143 | 1 | 1 |
| Yes | 4.9 | 0 | 0 (0.00-0.41) | 0 (undetermined) |
| Vaccination |  |  |  |  |
| No vaccination | 27.5 | 73 | 1 | 1 |
| One dose of vaccine** | 13.0 | 15 | 0.44 (0.25-0.76) | 0.45 (0.24-0.87) |
| Two doses of vaccine*** | 41.1 | 55 | 0.48 (0.34-0.68) | 0.51 (0.31-0.86) |
| Vaccine effectiveness one dose |  |  |  | 55% (13%-76%) |
| Vaccine effectiveness two doses |  |  |  | 49% (14%-69%) |

\*Adjusted for all variables shown in the Table and calendar week
\*\*Those with one dose are considered those who have received one dose of BNT162b2 mRNA COVID-19 vaccine more than 14 days prior.
\*\*\* Those with two doses are considered those who have received a second dose of BNT162b2 mRNA COVID-19 vaccine more than 7 days prior.

Table 3 displays the IRR of the association between characteristics of LTCF residents and severe COVID-19 related to B.1.351. Females were less likely to develop severe forms of COVID-19 disease compared to males (IRR = 0.35, 95%CI = 0.20-0.63). Vaccine effectiveness against severe forms of disease was estimated to be 86% (67-94).

**Table 3.** IRR of the association between characteristics of LTCF residents and severe COVID-19 related to B.1.351

|  | Person-years<br>(N=81.6) | N=52 | IRR<br>95% CI | IRRa*<br>95% CI |
| --- | --- | --- | --- | --- |
| Sex |  |  |  |  |
| Male | 18.7 | 23 | 1 | 1 |
| Female | 62.9 | 29 | 0.36 (0.21-0.62) | 0.35 (0.20-0.63) |
| Age (in years) |  |  |  |  |
| 55-84 | 24.5 | 13 | 1 | 1 |
| 85-94 | 47.2 | 36 | 1.29 (0.68-2.44) | 1.39 (0.72-2.68) |
| 94-104 | 9.9 | 3 | 0.58 (0.17-2.05) | 0.35 (0.09-1.30) |
| History of past SARS-CoV-2 infection |  |  |  |  |
| No | 76.7 | 52 | 1 | 1 |
| Yes | 4.9 | 0 | 0 (0.00-1.15) | 0 (undetermined) |
| Vaccination |  |  |  |  |
| No vaccination | 27.5 | 39 | 1 | 1 |
| One dose of vaccine | 13.0 | 2 | 0.12 (0.03-0.51) | 0.14 (0.03-0.68) |
| Two doses of vaccine | 41.1 | 11 | 0.17 (0.09-0.33) | 0.14 (0.06-0.33) |
| Vaccine effectiveness one dose** |  |  |  | 86% (32%-97%) |
| Vaccine effectiveness two doses*** |  |  |  | 86% (67%-94%) |

\*Adjusted for all variables shown in the Table and calendar week
\*\*Those with one dose are considered those who have received one dose of BNT162b2 mRNA COVID-19 vaccine more than 14 days prior.
\*\*\* Those with two doses are considered those who have received a second dose of BNT162b2 mRNA COVID-19 vaccine more than 7 days prior.

## Discussion

In this study, we have been able to show the protection of two doses of BNT162b2 mRNA vaccine against B.1.351 in LTCFs in eastern France. Vaccine effectiveness was estimated at 49% (14-69) against all forms of B.1.351 infection, and 86% (67-94) against severe forms of disease. These figures were lower when compared to those of a recent test-negative case-control study in Qatar, in which vaccine effectiveness against any and severe B.1.351 infection were 75% (71 to 79) and 97% (92 to 100), respectively.^16^ These differences may be explained by the older age of our study population (median age was 89 years) compared to that of the study in Qatar (median age of cases infected with B.1.351 was 33 years). Indeed, imunogenicity of the BNT162b2 mRNA vaccine appeared to be lower in adults aged 65-85 years compared to younger adults in clinical trials,^22^ and a study among LCTF residents in Denmark showed reduced effectiveness of BNT162b2 mRNA vaccine.^23^

We observed outbreaks of B.1.351 among residents of LTCFs, in spite of high vaccination coverage in LTCF D and E (in which 72% and 78% of residents had received two vaccine doses, respectively). These outbreaks were associated with severe forms of disease and deaths even among those who were fully vaccinated. Males were more likely to develop severe forms of disease as compared to females, as has been shown elsewhere.^24^ B.1.351 outbreaks and severe outcomes have recently been described among residents who had received two doses of BNT162b2 mRNA vaccine in a LTCF in eastern France, separate to those included in our study,^25^ and in a LTCF in Canada.^26^ However, none of these studies provided vaccine effectiveness estimates based on available data.

The main limitation of the study is the comparatively small amount of person-time participants contributed with one dose of vaccine only. This is likely the result of the short (three weeks) recommended interval between BNT162b2 mRNA vaccine doses at the time of the study. This makes the confidence intervals around the estimates of vaccine effectiveness associated with one dose of vaccine very broad. A further limitation was the lack of information available as part of the surveillance data beyond age, sex, and history of past infection of the residents. This prevented us from identifying whether some co-morbidities were associated with severe forms of disease.

The study was conducted shortly after BNT162b2 mRNA COVID-19 vaccine campaigns were initiated for LTCF residents in early 2021. As vaccine-related immunity is expected to decline more quickly in the older compared to the younger population,^27^ these breakthrough events four to six weeks after the second dose of vaccines advocate for the administration of a third dose in the fall prior to an expected increase in SARS-CoV-2 circulation this coming winter in the northern hemisphere.

Overall, our findings provide an important contribution to understanding the impact of B.1.351 lineage on the effectiveness of BNT162b2 mRNA COVID-19 vaccine in LTCFs. Vaccine effectiveness against B.1.351 was reduced, and we observed B.1.351 outbreaks with severe forms of disease among fully vaccinated individuals in two LTCFs. Our findings highlight the need to maintain SARS-CoV-2 surveillance in these high-risk settings beyond the current COVID-19 mass vaccination campaign.

## Data Availability

The data that support the findings of this study are available from the Societe de Pathologie Infectieuse de Langue Francaise. Restrictions apply to the availability of these data, which were used under authorized agreement for this study by the Societe de Pathologie Infectieuse de Langue Francaise, which ensures compliance with data protection regulations in France. Access to these data would therefore require prior authorization by the Societe de Pathologie Infectieuse de Langue Francaise.

## Contributors

BeL, CR and AF designed the investigation.

BeL, LT, CR and AF developed the study questionnaire.

BeL managed the data collection from LTCF.

LS oversaw the collection of the data and maintained the database.

LS, YM and AF performed the statistical analyses.

SW and BL organised the screening and sequencing of SARS-CoV-2 at the national level.

BeL, RG and AF drafted the first versions of the manuscript.

All authors critically reviewed and approved the final version of the manuscript.

## Data availability statement

The data that support the findings of this study are available from the Société de Pathologie Infectieuse de Langue Française. Restrictions apply to the availability of these data, which were used under authorized agreement for this study by the Société de Pathologie Infectieuse de Langue Française, which ensures compliance with data protection regulations in France. Access to these data would therefore require prior authorization by the Société de Pathologie Infectieuse de Langue Française.

## Declaration of interests

BeL reports travel funding from ViiV (2019) and Gilead (2020), outside the submitted work. All other authors have no competing interests.

## Acknowledgements

We would like to thank Professor Jérôme Salomon, Director General of Health in France, for facilitating this study. We would also like to thank all who were involved in this study: the Regional Health Agency of Grand-Est (Sandrine Guet, Jean-Pierre Gara, Olivier Dosso, Joël Restelli, Yves Le Balle, Franck Gerolt, Jean Le Moigne), the laboratory staff (Sebastien Fougnot, Jean Philippe Rault, Dominique Caby-Baer), the investigators from the long-term health care facilities (Mathilde Hemmer, Pauline Hannewald, Lola Antoine, Victoria Dennis, Camille Metzger, Elsa Remiatte, Alycia Layer, Emma Godard, Olivia Gouerec, Manon Nocus, Marie Di Sabatino, Ines Tabbone, Léa Willaume, Diego Caceres, Thibault Resch, Valentin Clément, Lucas Villemain, Quentin Garrouste, Aymeric Bernard), long-term care facility personnel (Dominique Huin, Laurence Zwahlen-Rolin, Caroline Guillotin, Céline Simonin, Sophie Hamel, Khadija Sghir, Jacqueline Genay, Marylin Guignard, Kathy Soriano, Emmanuelle Ulmer, Nathalie Muller, Emmanuelle Grossier, Christine Cornement, Marion Rosenau, Stéphanie Pietz, Magalie Jole, Céline Stickeir, Johnny Carpentier, Charles Sadoul, Jean-Maurice Demurger, Denis Gerber, Jean-Marc Roth, Arnaud Masson), and Laura Schaeffer who helped with set up of the questionnaire and the database.

## Funding

This study has benefited from financial support from the French Ministry of Health and Solidarity.

## Notes

### Competing Interest Statement

Benjamin Lefevre reports travel funding from ViiV (2019) and Gilead (2020), outside the submitted work.
All other authors have no competing interests.

### Author Declarations

In France, surveillance of COVID-19 testing and COVID-19 vaccination data in LTCFs is considered as public health surveillance which does not require formal ethical review. The pseudonymized data collection by medical personnel at the LTCFs and analysis by the authors have been performed under the legal responsibility of the Societe de Pathologie Infectieuse de Langue Francaise, which ensured compliance with data protection regulations in France.

